# Ex Vivo Drug Responses and Molecular Profiles of 597 Pediatric Acute Lymphoblastic Leukemia Patients

**DOI:** 10.1101/2024.12.17.24319138

**Authors:** Anna Pia Enblad, Olga Krali, Henrik Gezelius, Anders Lundmark, Kristin Blom, Claes Andersson, Josefine Palle, Britt-Marie Frost, Samppa Ryhänen, Trond Flægstad, Ólafur G Jónsson, Kjeld Schmiegelow, Mats Heyman, Arja Harila, Peter Nygren, Rolf Larsson, Gudmar Lönnerholm, Jessica Nordlund

**Author notes:** **Corresponding author:** Dr. Jessica Nordlund, Biomedical Centre (BMC), Husargatan 3, Box 1432, SE-75144 Uppsala, Sweden. **Email addresses of co-authors:** Anna Pia Enblad, Olga Krali, Henrik Gezelius, Anders Lundmark, Kristin Blom, Claes Andersson, Josefine Palle, Britt-Marie Frost, Samppa Ryhänen, Trond Flægstad, Ólafur G Jónsson, Kjeld Schmiegelow, Mats Heyman, Arja Harila, Peter Nygren, Rolf Larsson, Gudmar Lönnerholm.

## Abstract

*Ex vivo* drug response profiling is emerging as a valuable tool for identifying drug resistance mechanisms and advancing precision medicine in hematological cancers. However, the functional impact of dysregulation of the epigenome and transcriptome in this context remains poorly understood. In this study, we combined *ex vivo* drug sensitivity profiling with transcriptomic and epigenomic analyses in diagnostic samples from 597 pediatric B-cell precursor acute lymphoblastic leukemia (BCP-ALL) patients. *Ex vivo* resistance to antimetabolites (e.g., cytarabine, thioguanine), glucocorticoids (e.g., dexamethasone, prednisolone), and doxorubicin was independently associated with reduced relapse-free survival (RFS, p < 0.05). Molecular profiling identified pre-treatment DNA methylation and gene expression patterns distinguishing resistant from sensitive cases, revealing key drug resistance signatures. These included aberrant expression of genes related to heme metabolism (e.g. *ATPV06A*) and *KRAS* signaling (e.g. *ABCB1*). Notably, we also observed atypical expression of genes usually restricted to T-cells and other immune cells (e.g., *ITK*) in resistant BCP-ALL cells. Our findings demonstrate that *ex vivo* drug response patterns are predictive of clinical outcomes and reflect intrinsic molecular states associated with drug tolerance. This integrative multi-omics approach highlights potential therapeutic targets and underscores the value of functional precision medicine in identifying treatment vulnerabilities in pediatric ALL.

**Key points:**

- *Ex vivo* drug response profiling provides predictors of outcome that complement standard clinical risk stratification, identifying high-risk phenotypes not captured by traditional methods.
- Pre-treatment transcriptional and epigenetic profiles reveal resistance-primed molecular states detectable at diagnosis, mechanisms driving treatment resistance and offer potential therapeutic targets to counteract drug tolerance.

## Introduction

Therapeutic advances in pediatric acute lymphoblastic leukemia (ALL) have significantly improved patient outcomes, with modern treatment protocols leveraging risk stratification based on clinical presentation, genetic markers, and minimal residual disease (MRD).^1,2^ However, 10-20% of patients still relapse or develop resistant disease^3^, including a notable proportion of those initially classified as low-risk.^2,4^ Moreover, while most patients are cured, the intense treatment regimens often result in significant short- and long-term toxicities.^5–8^ Evidence indicates many standard-risk patients could benefit from less intensive chemotherapy, highlighting the limitations of current risk stratification methods.^9,10^ Emerging immune therapies, such as blinatumomab and CAR-T cells, have shown promise in the relapsed and refractory settings^11,12^ and are increasingly explored as alternatives or supplements in ALL therapy.^13^ However, the results of such options are affected by the remaining tumor burden^14,15^, thus initial response to chemotherapeutic agents is key for achieving lasting remission. Addressing these challenges requires a deeper understanding of the mechanisms underlying treatment response and resistance.

*Ex vivo* drug screening evaluates the response of patient-derived leukemia cells to therapeutic agents *in vitro*, offering a direct measure of cellular drug efficacy. This approach has potential as a predictor of clinical outcomes in pediatric ALL.^16–19^ Combining *ex vivo* drug screening with omics-based assays has demonstrated the clinical feasibility of “functional precision medicine”, enabling the identification of therapeutic options across various malignancies, including pediatric ALL.^20–23^ However, these efforts have primarily focused on relapsed, treatment-refractory, or high-risk cases. While whole-genome sequencing is a powerful tool for identifying genetic variants associated with disease progression^24^, it fails to capture the full spectrum of regulatory mechanisms and cellular processes driving drug resistance. ^25^ Among these, epigenetic alterations, which are increasingly recognized a contributors to drug resistance ^26–28^, remain under explored. Promising work has linked epigenetic profiles to *ex vivo* drug resistance in pediatric ALL.^25,29^ However, these studies have largely focused on single-drug responses, which do not reflect the multi-drug chemotherapy regimens used in clinical practice. Expanding these investigations, particularly in diagnostic samples and multi-drug contexts will be essential to evaluate the future utility of such approaches for functional precision medicine strategies.

In this study, we performed *ex vivo* drug response profiling on a Nordic cohort of diagnostic samples from 597 pediatric BCP-ALL patients, integrating the responses to 10 therapeutic agents with transcriptomic and epigenomic data. This approach provided clinically relevant risk stratification information and identified molecular markers and pathways associated with drug resistance. These findings underscore the potential of integrative strategies to uncover pre-treatment molecular states that predispose patients to drug resistance.

## Methods

### Patient cohort

The study consisted of 597 children (1–18 years of age) diagnosed with BCP-ALL between 1992 and 2008 at Nordic centers for pediatric oncology. Patients were treated according to the Nordic Society of Pediatric Hematology and Oncology (NOPHO) treatment protocols (NOPHO ALL-92 or NOPHO ALL-2000) and were stratified into risk groups at the time of diagnosis: standard risk (SR), intermediate risk (IR) or high risk (HR) according to established criteria^30^, including age, white blood cell count at diagnosis, extramedullary involvement and molecular genetic subtype. The patients and/or guardians provided informed consent. The study was approved by the Regional Ethical Review Board in Uppsala, Sweden, and was conducted according to the guidelines of the Declaration of Helsinki.

### Clinical outcome

Follow up data and clinical outcome was reported by the participating clinics to the NOPHO registry at the Childhood Cancer Research Unit in Stockholm. Complete remission (CR) was defined as less than 5% leukemic blasts in a representative bone marrow sample and no other manifestation of leukemia. Relapse was defined as more than 5% blast cells in bone marrow and/or manifestation of leukemia at any other location. Relapse-free survival (RFS) was defined as the time from diagnosis to leukemic relapse at any location and overall survival (OS) was defined as the time from diagnosis to death by any cause.

### Sample processing and *ex vivo* drug screening procedure

Bone marrow samples were collected at diagnosis in heparinized glass tubes and sent to Uppsala, Sweden for analysis. *Ex vivo* drug responses were assessed by the fluorometric microculture cytotoxicity assay (FMCA)^31^ and the experimental procedures including handling of ALL samples follows Frost et al.^17^ In brief, FMCA generates a survival index (SI%) for each drug per patient, where a high numerical SI% indicates *ex vivo* resistance to the drug. Ten drugs were tested at concentrations chosen to give high SI% variability between samples, based on previous work.^32^ The drugs represent different drug classes, including compounds used in the NOPHO ALL-92 and ALL-2000 treatment protocols (dexamethasone, prednisolone, doxorubicin, vincristine, L-asparaginase, cytarabine, thioguanine) as well as three additional drugs (mitoxantrone, amsacrine, etoposide). Drugs, concentrations, and the number of patients screened per drug summarized in Supplemental table 1.

### DNA methylation and RNA sequencing data

DNA and RNA were extracted as previously described from drug-naïve diagnostic samples.^33^ Genome-wide DNA methylation (DNAm) data were generated with the Infinium HumMeth450K BeadChip assay (450k array, n = 382 patients) or the MethylationEPIC v2.0 BeadChip assay (EPIC array, n = 55 patients).^34,35^ RNA-seq data (GEX) was generated for 119 patients.^34,36^ The computational approaches to analyze the DNAm and GEX datasets are described in detail previously^34,35^ and further summarized in the Supplemental methods. After filtering and normalization, a total of 372,264 CpG sites (DNAm) and 15,019 genes (GEX) were available downstream analyses.

### Molecular genetic subtype revision

At the time of ALL diagnosis, patients were divided into subtype groups based on immunophenotype and recurrent genetic aberrations as established by FISH, PCR, or G-band karyotyping.^30^ Since the patients were diagnosed before the introduction of high throughput sequencing techniques, complete information was unavailable for 224 patients (B-other, unknown, or uncertain subtype). We previously performed additional molecular analysis and machine learning-based classification^34^, resulting in subtypes re-classification of 106 patients to the current International Consensus Classification.^37^ A summary of molecular subtypes is provided in Table 1.

**Table 1.** Patient cohort characteristics.

| Study cohort characteristics |  | n(%) |
| --- | --- | --- |
| All patients |  | 597 (100.0) |
| Mean age (SD) |  | 6.1 (4.1) |
| Age group (years) |  |  |
| <5 |  | 314 (52.6) |
| 5-10 |  | 186 (31.2) |
| 11-14 |  | 68 (11.4) |
| ≥15 |  | 29 (4.9) |
| Sex |  |  |
| Female |  | 274 (45.9) |
| Male |  | 323 (54.1) |
| Risk group |  |  |
| Standard |  | 194 (32.5) |
| Intermediate |  | 266 (44.6) |
| High |  | 137 (22.9) |
| Subtype |  |  |
| High hyperdiploid |  | 215 (36.0) |
| ETV6::RUNX1 |  | 136 (22.8) |
| B-other |  | 118 (19.8) |
| PAX5alt |  | 26 (4.4) |
| TCF3::PBX1 |  | 22 (3.7) |
| KMT2A-rearranged |  | 17 (2.8) |
| DUX4-rearranged |  | 16 (2.7) |
| ZNF384-rearranged |  | 9 (1.5) |
| iAMP21 |  | 8 (1.3) |
| BCR::ABL1-like |  | 8 (1.3) |
| ETV6::RUNX1-like |  | 8 (1.3) |
| BCR::ABL1 |  | 7 (1.2) |
| MEF2D-rearranged |  | 4 (0.7) |
| PAX5 p.Pro80Arg |  | 3 (0.5) |
| Primary event |  |  |
| Complete remission 1 |  | 437 (73.2) |
| Relapse |  | 134 (22.4) |
| Death in complete remission 1 |  | 10 (1.7) |
| Induction failure |  | 7 (1.2) |
| Secondary malignant neoplasm |  | 7 (1.2) |
| Resistant disease |  | 2 (0.3) |
| Status |  |  |
| Alive |  | 515 (86.3) |
| Dead |  | 82 (13.7) |

### Bioinformatics and statistical analysis

Data processing and statistical analyses were performed using RStudio (RStudio 2022.07.2 “Spotted Wakerobin” and RStudio 2023.03.15 “Shortstop Beagle”) or Python. For all analyses, a p-value < 0.05 was considered statistically significant, if not otherwise specified.

Survival analysis was performed using the R package *ggsurvfit* to create Kaplan-Meier curves. Log-rank tests were used to test for differences in survival between groups. Multivariate Cox regression was performed using the R package *survival*, with *ex vivo* drug response group, risk group and sex as covariates.

Unsupervised hierarchical clustering was performed using the Python package *scipy*. Missing values for *ex vivo* drug responses were imputed patient-wise with the most frequent response category for each patient (sensitive, intermediate or resistant). Spearman’s rank correlation coefficient was performed to assess the relationship between drug responses (SI%).

Differentially methylated CpG sites (DMCs) were determined in the R package *limma*^38^ using a FDR adjusted p-value of < 0.01 and mean absolute delta beta-value > 0.15 between groups. Differentially expressed genes (DEGs) were determined using *DeSeq2*^39^ using FDR adjusted p-value < 0.05 and absolute mean log2FC > 1 between groups. ALL molecular subtype was adjusted for when testing for DMCs and DEGs. DMCs were annotated to genes using GREAT 4.0.4.^40^ Gene set analysis was performed using the MSigDB Hallmark^41^ and gene ontology biological processes.^42^

### Single-cell analysis

Bone marrow samples from four BCP-ALL patients diagnosed with *ETV6::RUNX1-*positive pre-B ALL were selected for single-cell RNA-sequencing (scRNA-seq). Vital frozen ampules after Ficoll gradient separation were collected from NOPHO Leukemia Biobank in Uppsala, Sweden. The cells were thawed and seeded to microwell plates. Each of the wells were incubated with 0.45 vol% DMSO for 72 hours before collection. Cells were harvested by gentle centrifugation (300 rpm 10 minutes) and washed twice with PBS. ScRNA-seq libraries were generated using the 10x Genomics FLEX kit following the manufacturer’s protocol. The libraries were sequenced on an Illumina NovaSeq X plus 10B flowcell with the following read setup: 28 bp -10 bp-10 bp-90 bp. Cellranger 8.0.1 was used to demultiplex single cell sequence data into UMI count matrices. Downstream analysis of expression data was performed in R 4.4.1 using *Seurat 5.1.0*.^43^ Cells were called using the inflection point determined by *barcodeRanks* from the package *DropletUtils* 1.25.2.^44^ Remaining cells with a mitochondrial gene percentage greater than ten percent were removed. Cell types were assigned using *BoneMarrowMap 0.1.0.* ^45^

## Results

### Patient characteristics

A total of 597 children with BCP-ALL, treated according to the NOPHO ALL-1992 (45.6%, n = 272) or ALL-2000 (54.4%, n = 325) protocols, were included in the study (Table 1). Leukemic relapse occurred in 134 patients, with proportions varying between risk groups; 21% of SR patients, 18% of IR patients and 32% of HR patients relapsed. During follow-up, 82 patients died; 61 after relapse, ten in complete remission, seven following induction failure, three following secondary malignant neoplasm and one due to resistant disease. The outcomes of the selection of patients in this study is similar to what was reported overall for all patients treated on NOPHO ALL-1992 and -2000.^30^ For most of the drugs ∼90% of the patients were screened for *ex vivo* response, however the proportion was lower for cytarabine (73%), mitoxantrone (72%), and L-asparaginase (42%) (Supplemental table 1).

### Ex vivo drug resistance is an independent predictor of clinical outcome

Patients were stratified into three *ex vivo* drug response categories (sensitive, intermediate, and resistant) based on tertiles of surviving cells (SI%) for each drug (Figure 1A). The proportions of samples in each tertile group were similar across countries of origin, indicating that transportation time did not impact the *ex vivo* screening results (Supplemental table 2).

**Figure 1.**
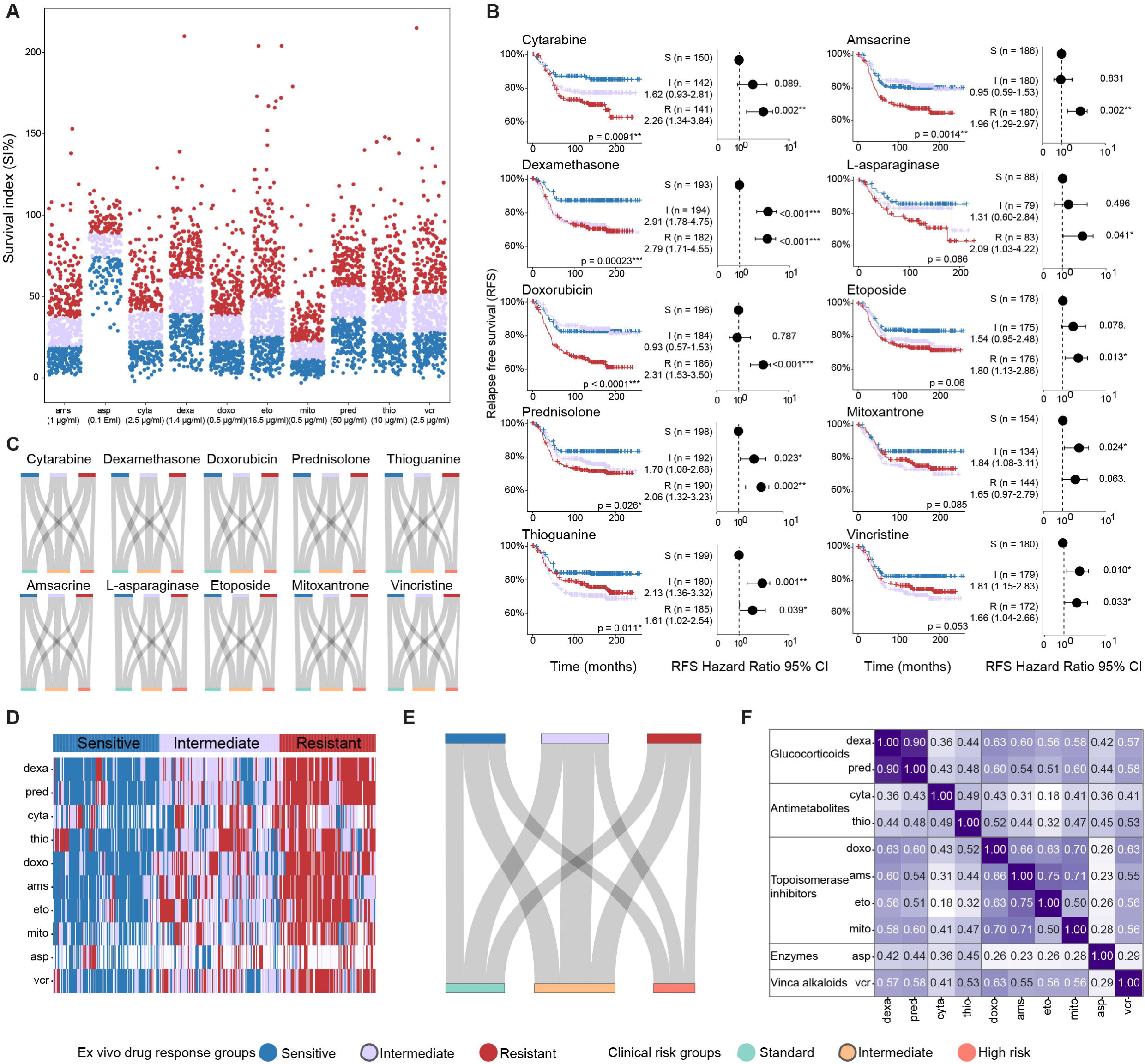
Ex-vivo drug response and clinical outcome in pediatric BCP-ALL patients. A) SI% distribution (y-axis) per drug (x-axis). The patients are color-coded by their *ex vivo* drug response group (sensitive, intermediate, resistant). B) Kaplan Meier curves depicting relapse free survival, with patients stratified by the *ex vivo* drug response groups (sensitive, intermediate or resistant), p-values calculated using log-rank test. Forest plots showing hazard ratio and 95% confidence interval for relapse in groups with different *ex vivo* drug response, adjusted for sex and risk group. C) Relationship between the distribution of patients in different *ex vivo* drug response groups and the different clinical risk groups. D) Unsupervised hierarchical clustering of patients based on *ex vivo* drug response to the ten individual drugs, rendering three main clusters deemed multi-drug sensitive, intermediate and resistant. E) Relationship between the distribution of patients in different levels of *ex vivo* multi-drug response and the different clinical risk groups. F) Correlation plot displaying the Spearman’s rank correlation coefficient pairwise between different drugs, to assess the relationship between drug responses. Dexa = dexamethasone, pred = prednisolone, cyta = cytarabine, thio = thioguanine, doxo = doxorubicin, ams = amsacrine, eto = etoposide, mito = mitoxantrone, asp = L-asparaginase, vcr = vincristine.

Patients in the *ex vivo* sensitive group demonstrated superior relapse-free survival (RFS) compared to those in the resistant group for a majority of the drugs, while the outcomes for the intermediate group varied by drug (log rank test p < 0.01; Figure 1B). Notably, *ex vivo* responses to dexamethasone, doxorubicin, and thioguanine were significantly associated with overall survival (OS, log rank test p < 0.01, Supplemental Figure 1). These associations remained significant in multivariate Cox regression analyses, adjusted for clinical risk group and sex (Figure 1B, Supplemental Figure 1).

Next, we assessed the distribution of clinical risk groups within the three-tiered *ex vivo* response categories. Surprisingly, there was no enrichment of high-risk patients in the *ex vivo* drug-resistant groups. Instead, resistant patients were evenly distributed across all clinical risk groups (Figure 1C). When responses to all 10 drugs were considered collectively, a pattern of *ex vivo* multidrug resistance emerged (Figure 1D). Similar to single-drug observations, multidrug resistance was not associated with clinical risk groups (Figure 1E), but was strongly correlated with inferior clinical outcomes, including RFS and OS (Supplemental Figure 2).

Additionally, patients displayed similar *ex vivo* responses to drugs with shared mechanisms of action (Figure 1F), suggesting that intrinsic cellular properties influence drug responses based on their mechanisms. Together, these findings demonstrate that *ex vivo* drug responses reflect *in vivo* drug efficacy and may complement traditional clinical risk stratification methods.

### Pre-treatment molecular profiles are associated with ex vivo response

We next investigated molecular profiles of drug-naïve diagnostic samples in relation to *ex vivo* responses. To focus on the most pronounced differences, the resistant and sensitive groups were compared, excluding samples from the intermediate response group from this analysis. This analysis identified 1,779 unique DMCs and 1,158 unique DEGs between the sensitive and resistant groups across the 10 drugs (Figure 2A-D, Supplemental Tables 3 and 4). Notably, the most pronounced differences in molecular profiles were observed in cells resistant to the glucocorticoids and topoisomerase II inhibitors, while few DMCs or DEGs were correlated with response to antimetabolites and L-asparaginase.

**Figure 2.**
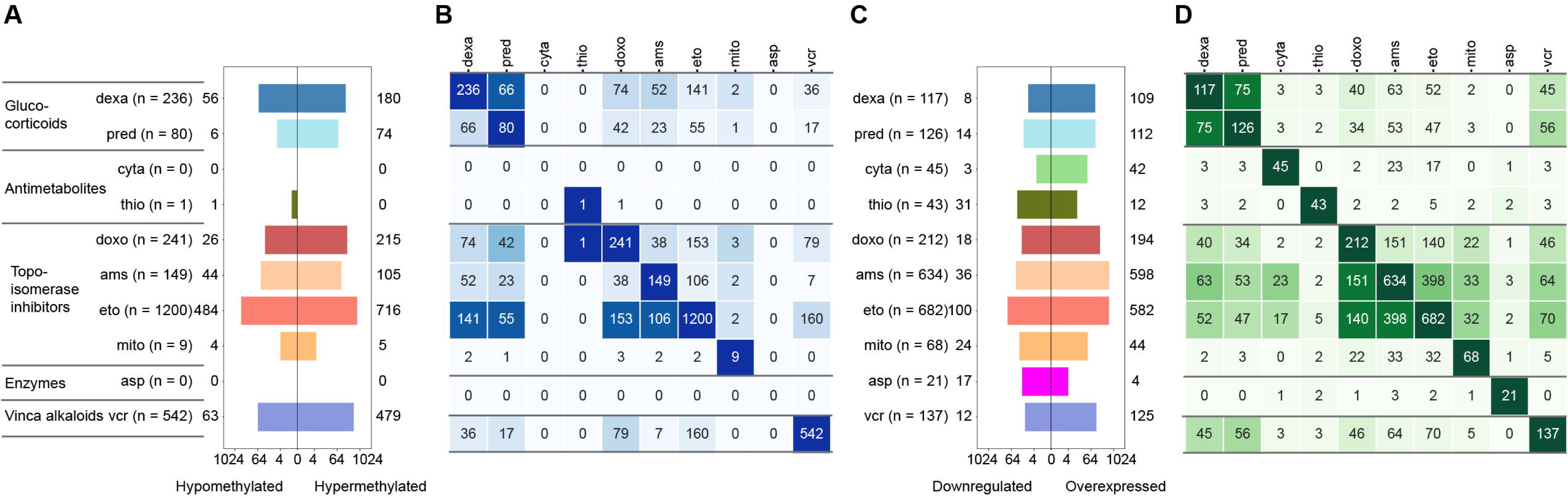
Molecular profiles related to ex vivo drug responses. A) Numbers of differentially methylated CpG sites (DMCs) per drug, with total numbers indicated in parenthesis and number of hypo- and hypermethylated CpG sites shown left and right of the bar graph. B) Pairwise comparison between drugs displaying the number of overlapping DMCs between drugs, colored according to percentage of DMCs overlapping. C) Numbers of differentially expressed genes (DEGs) per drug, with total numbers indicated in parenthesis and number of downregulated and overexpressed genes shown left and right of the bar graph. D) Pairwise comparison between drugs displaying the number of overlapping DEGs between drugs, colored according to percentage of DEGs overlapping.

The number of drug-related DMCs varied by drug, ranging from none for L-asparaginase and cytarabine to 1,200 for etoposide (Figure 2A). These DMCs were enriched in low-CpG content, open sea regions (Supplemental figure 3). Notably, 24% of DMCs overlapped between at least two drugs, with the greatest overlap observed among drugs with similar mechanisms of action (Figure 2B). For example, 34 DMCs were shared by the three topoisomerase II inhibitors (doxorubicin, etoposide, amacrine) and 66 by the glucocorticoids (dexamethasone, prednisolone).

Similarly, the number of DEGs varied by drug, from 21 for L-asparaginase to 682 for etoposide (Figure 2C). Up to 49% of DEGs were shared between drugs, with notable overlaps observed for glucocorticoids and topoisomerase II inhibitors (Figure 2D). For instance, 398 DEGs were shared between etoposide and amsacrine, 75 between dexamethasone and prednisolone, and a significant overlap was also found between vincristine, glucocorticoids, and topoisomerase II inhibitors (Figure 2D). These findings suggest distinct molecular profiles in drug-naïve diagnostic samples, with shared, as well as drug-specific signatures.

To explore the interplay between DNA methylation and gene expression, we focused on 47 genes with DMCs and DEGs linked to the same drug (Supplemental table 5). Most were associated with resistance to etoposide, amsacrine, dexamethasone and doxorubicin. Notably, three genes that were hypomethylated and upregulated in the resistant samples where *ABCB1*, encoding the multidrug resistance-associated ABC transporter P-gp1; *ADHFE1*, a MYC-related oncogene linked to cell survival; and *CXCL2* that has been associated with increased leukemic cell survival and poor prognosis in ALL and other cancer*s.*^46–48^ These finding further emphasize the potential mechanistic link between epigenetic regulation and gene expression in the context of intrinsic cellular resistance.

### Pathways priming resistance

To identify pathways influencing drug response, we performed gene set analysis, for the genes mapped to DMCs and DEGs independently. Overlap in pathways was observed within and across drug classes and data modality (Figure 3A, Supplemental Table 6). Among the top-ranking pathways was upregulation of KRAS signaling (including e.g. *ABCB1, GS02)*, identified for cells resistant to topoisomerase II inhibitors, as has been previously shown for etoposide^49^ and doxorubicin.^50^ Additionally, heme metabolism was affected, with for example upregulation of *OSBP2* and *ATP6V0A1* in resistant patients (Figure 3B). These two genes have previously been linked to chemoresistance.^51,52^ Immune response-related pathways were also upregulated in patients resistant to topoisomerase II inhibitors and vincristine. Increased inflammatory activity, previously tied to chemotherapy resistance and poor outcomes in leukemia, supports the relevance of these findings.^53^

**Figure 3.**
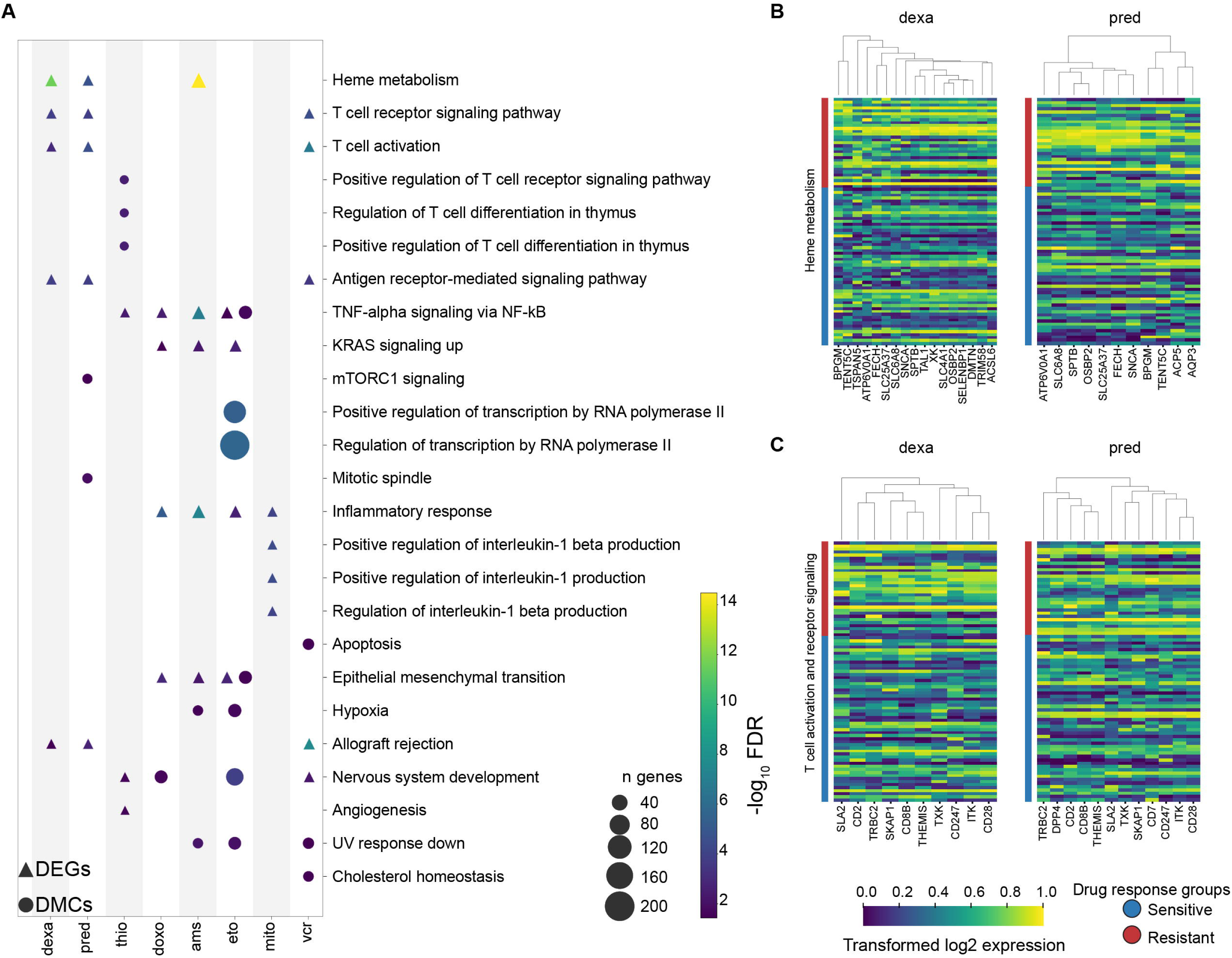
Pathways priming resistance in BPC-ALL. A) The top three most significantly enriched pathways per drug (lowest FDR adjusted p-value) and, in case of the same pathways being found for other drugs, this is also displayed, even if it was not in the top three most significantly enriched pathways for that drug. Round shape indicates that the pathway was related DMCs and triangle shape that the pathway was associated with genes related to DEGs. Size indicates how many genes were enriched to each pathway, colored according to the -log_10_ FDR score. B) Heatmaps showing the different expression levels of genes (log2 expression) related to heme metabolism and C) T-cell pathways (bottom), between glucocorticoid (dexamethasone and prednisolone) resistant and sensitive patients.

Interestingly, pathways related to T-cell linage were enriched in the samples resistant to four important anti-leukemia drugs (dexamethasone, prednisolone, thioguanine and vincristine). The associated genes were either upregulated or hypomethylated in resistant patients and included genes typically expressed by non-B-cell lineage immune cells (T-cells and NK-cells), such as *ITK* and *TXK* (Figure 3C).

### Validation by single-cell RNA-sequencing

To confirm that the DEGs identified in the drug-related pathways were intrinsic to leukemic cells and not artifacts due to other cell types present in bulk samples, we performed single-cell RNA sequencing (scRNA-seq) on diagnostic bone marrow samples from four ALL patients (Figure 4A, patient characteristics in Supplemental table 7). While most cells in these samples were annotated to the B-cells linage, consistent with the leukemic population, also additional cell types could be detected (Figure 4B). Still, key genes from our pathway analysis, including *ABCB1* and *G0S2* (KRAS pathway)*, OSBP2* and *ATP6V0A1* (heme metabolism), *ITK* and *TXK* (T-cell related genes) were expressed in the B-lineage leukemic cells (Figure 4C). Notably, we observed variability in gene expression across the cells from the four patients, reflecting heterogeneity within the leukemic cell population. These results support that aberrant gene expression identified in bulk analyses originates from BCP-ALL cells rather than contamination by non-leukemic cells, strengthening the relevance of our findings.

**Figure 4.**
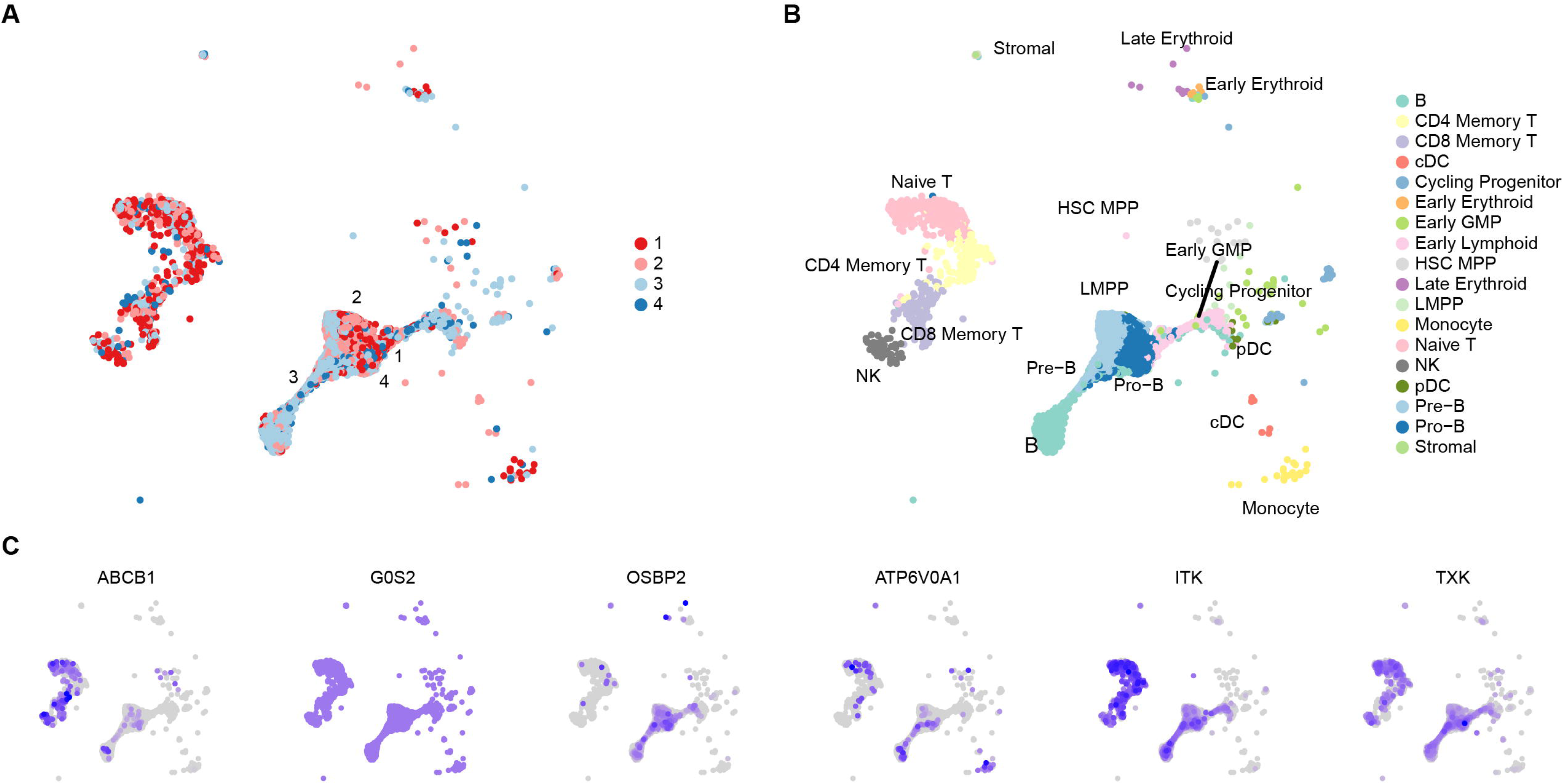
Single-cell (sc) profiling of pediatric BCP-ALL samples. A) sc-UMAP plot color-coded by patient ID. B) sc-UMAP plot colored by cell linage. C) UMAP plots demonstrating gene-specific expression for genes involved in KRAS *(ABCB1, G0S2*), heme (*OSBP2*, *ATP6V0A1*) and T-cell related pathways (*ITK* and *TXK*).

## Discussion

In this study, we combined *ex vivo* drug screening with molecular profiling of drug-naïve samples from 597 pediatric BCP-ALL patients. Our findings highlight two key insights: first, *ex vivo* drug responses serve as robust predictors of clinical outcomes; second, molecular signatures identified in diagnostic samples reveal intrinsic cellular states associated with increased tolerance or resistance to treatment. Together, these observations strengthen the case for functional precision medicine in treating ALL.

Early identification of patients resistant to conventional therapies through *ex vivo* approaches could potentially enable the upfront recognition of high-risk cellular phenotypes and guide the selection of alternative therapeutic strategies. Our findings provide orthogonal evidence supporting the potential benefit integrating such approaches for ALL. Consistent with previous studies, out data suggest that these types of functional assays offer additional information that can cooperate with genetic analyses risk grouping.^20–22,54–56^ Further, our observation of patients resistant to one drug were more likely to exhibit resistance across multiple drugs with different mechanisms, underscoring the relevance of addressing multidrug resistance in ALL.^57,58^

The idea of drug response-associated states that exists prior to any chemotherapy have been seen before.^59^ Using bulk molecular profiling (RNA-seq and DNAm arrays) in drug-naïve samples, we identified putative molecular states, priming cells for resistance, highlighting several candidate genes and pathways for future investigation. Among the genes we identified was *ABCB1*, encoding P-gp1, an efflux transporter that mediates the efflux of diverse drugs from cancer cells.^60,61^ Notably, we found *ABCB1* to be variably expressed in BCP-ALL cells, with significantly higher levels in resistance-primed patients. These findings prioritize *ABC* modulators, including specifically modulators of *ABCB1*^62^ for further exploration as a potential treatment in combination with standard chemotherapeutic drugs to augment therapeutic efficacy in BCP-ALL.

We also found three pathways of particular interest: KRAS signaling, heme metabolism and expression of T-cell related genes. Mutations affecting the Ras pathway, including KRAS mutations, are common in BCP-ALL and has previously been linked to poor outcome.^63–65^ Herein, we found that KRAS activation was correlated with resistance to topoisomerase II inhibitors. Although inhibiting KRAS has long been deemed difficult, new approaches for targeting KRAS in cancer therapy are emerging, and our results further support continuing this avenue for ALL.^66,67^ The importance of heme metabolism in chemotherapy resistance has been noted in AML^68^ as well as other non-hematological cancer types,^69,70^ but to our knowledge has not been observed previously in BCP-ALL. Upregulation of *ATP6V0A1*, which encodes a subunit of a vacuolar ATPase (V-ATPase) proton pump, was a particularly interesting observation as it has functionally established role in chemotherapy resistance,^52,71^ where targeted inhibitors exist.^72,73^ Finally, we observed increased expression of T-cell lineage genes in patients *ex vivo* resistant to the drugs that are central elements of ALL treatment (glucocorticoids, vincristine and thioguanine). Aberrant T-lineage cell surface marker expression in B-cell malignancies has previously been linked to poor outcome,^74^ but the expression of T-cell lineage genes in BCP-ALL cells and the relationship to drug response has not been described. One gene in particular, *ITK,* was upregulated in *ex vivo* resistant patients. Ibrutinib, a *BTK* inhibitor effective in chronic lymphocytic leukemia^75^ and in combination with dasatinib for of CNS-infiltrating BCP-ALL^76^, has also shown promising results in inhibiting *ITK*^77^. Our results broaden the potential scope of *BTK* inhibition for treatment of BCP-ALL.

Challenges faced by our approach include sample handling and cell viability.^78^ Furthermore, our study primarily assessed responses to individual drugs, whereas BCP-ALL treatment involves complex, multi-drug regimens administered over extended timeframes. These regimens may produce synergistic or antagonistic effects that single-drug assays cannot fully replicate. However, advancements in functional assay technologies are making high-throughput platforms increasingly robust and scalable, enabling the simultaneous screening of hundreds of drugs in various combinations.^79–81^ These developments hold promise for addressing the limitations of single-drug analyses in future studies.

Here, our bulk analysis approach may additionally obscure heterogeneity within leukemic cell populations by averaging signals across the sample, which likely includes both leukemic and normal cells as well as sub-populations with distinct molecular and genetic diversity^82^. Single-cell technologies combined with *ex vivo* drug screening can address some of these limitations by allowing the investigation of drug responses at the resolution of individual cells.^83–85^ Moreover, sequential drug treatment studies have demonstrated promise in blocking non-genetic cancer evolution, underscoring the importance of capturing dynamic cellular responses over time.^86^ Integrating these advanced approaches with multi-omics profiling in future studies could provide deeper insights into treatment resistance mechanisms and therapeutic vulnerabilities that remain beyond the scope of bulk analyses. However, it is important to note that in our study, we identified molecular signatures in bulk samples that were also observed at the single-cell level, affirming the relevance and robustness of our findings despite the limitations of bulk approaches.

In conclusion, our findings demonstrate that BCP-ALL patient cells exhibit distinct molecular signatures associated with drug resistance, highlighting pre-treatment mechanisms that may prime cells for therapy failure. These results underscore the value of integrating *ex vivo* drug screening with molecular profiling to identify resistance mechanisms and uncover potential therapeutic targets, paving the way for improved treatment strategies and outcomes.

## Data Availability

The DNA methylation data were available from their original studies and the metadata are available at the Gene Expression Omnibus (GEO) under accession number GSE49031, and under controlled access via 10.17044/scilifelab.22303531 and 10.17044/scilifelab.26096371 (https://figshare.scilifelab.se/). The gene expression data are available under GSE227832. Data requests may be submitted to Jessica Nordlund.

## Authorship Contributions

G.L., and J.N. conceived the study. A.P.E, O.K., A.L, H.G, J.N and G.L. analyzed the data. A.P.E, O.K., A.L. and J.N. designed the figures. K.B. and H.G. performed experiments. J.P., B.M.F., S.R., T.F., O.G.J., K.S., M.H., A.H. and G.L. provided clinical material, patient data and clinical expertise. P.N. and R.L. established methods. K.B., C.A., P.N. and R.L. provided *ex vivo* drug screening expertise. A.P.E., O.K., and J.N. wrote the manuscript. All authors read and approved the final version of the manuscript.

## Acknowledgements

We thank Martin Åberg for cell preparations for the scRNA-seq analysis. This work was supported by grants from the Swedish Research Council (2019-01976 to JN), the Swedish Cancer Society (CAN2022-2395 to JN), the Swedish Childhood Cancer Fund (PR2022-0082 and HFT2023-0011 to JN, TJ2020-0039 to AH), and the Göran Gustafssons Foundation (to JN). The genomics data was generated with assistance from the SciLifeLab National Genomics Infrastructure, SNP&SEQ Technology Platform, which is funded by the Swedish Research Council and the Knut and Alice Wallenberg Foundation. Computational resources were provided by the Swedish National Infrastructure for Computing (SNIC) and National Academic Infrastructure for Supercomputing in Sweden (NAISS).

