## Supplementary Table 1 for "Ex Vivo Drug Responses and Molecular Profiles of 597 Pediatric Acute Lymphoblastic Leukemia Patients"

| **Drug** | **Nr of patients ex-vivo drug screened (%)** | **Concentration** |
|---|---|---|
| Amsacrine | 546 (91) | 1 µg/ml |
| L-asparaginase | 250 (42) | 0.1 Eml |
| Cytarabine | 433 (73) | 2.5 µg/ml |
| Dexamethasone | 569 (95) | 1.4 µg/ml |
| Doxorubicin | 566 (95) | 0.5 µg/ml |
| Etoposide | 529 (89) | 16.5 µg/ml |
| Mitoxantrone | 432 (72) | 0.5 µg/ml |
| Prednisolone | 580 (97) | 50 µg/ml |
| Thioguanine | 564 (94) | 10 µg/ml |
| Vincristine | 531 (89) | 2.5 µg/ml |
