## Supplementary Figure 1 for "Ex Vivo Drug Responses and Molecular Profiles of 597 Pediatric Acute Lymphoblastic Leukemia Patients"

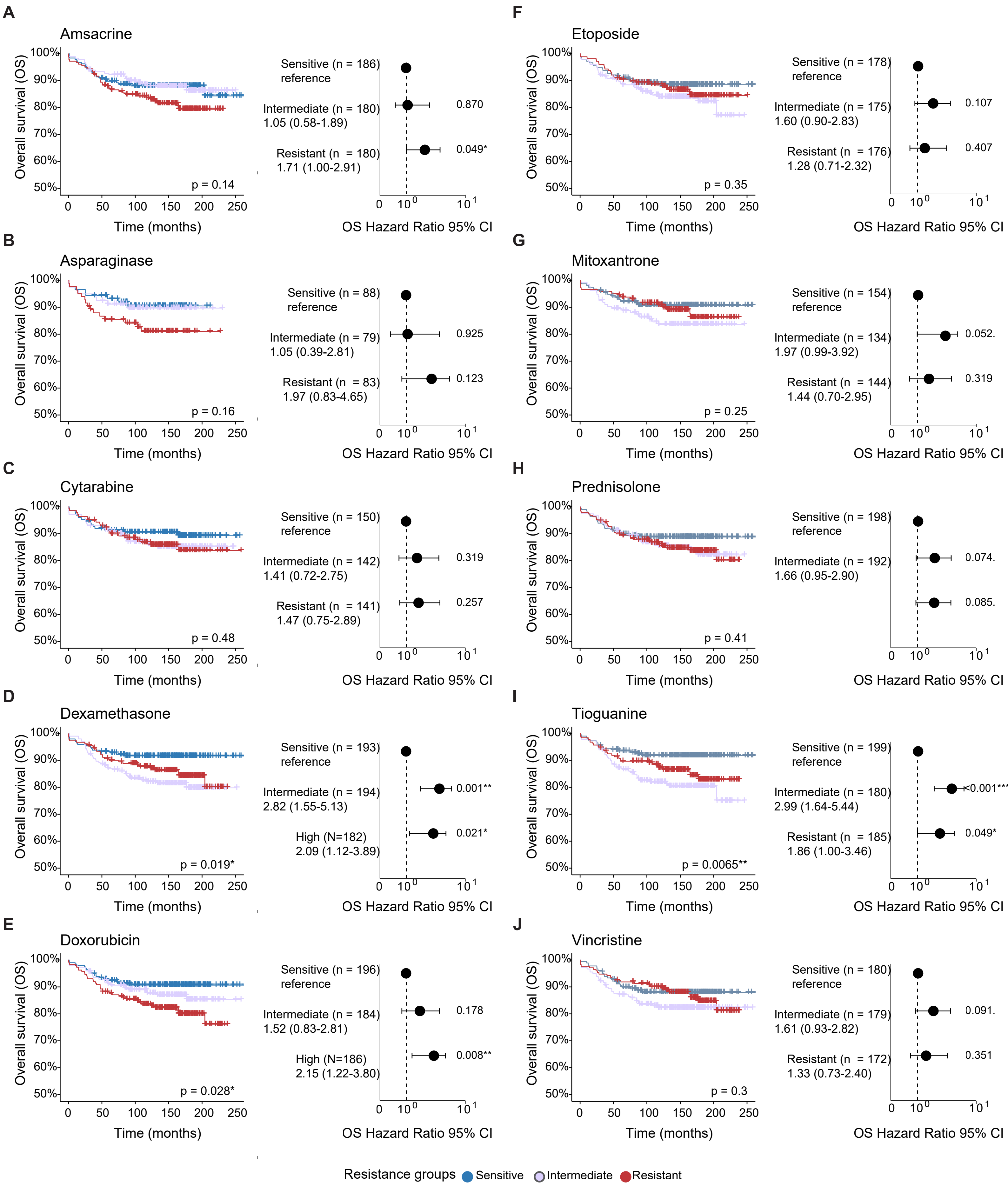


**Supplemental Figure 1. Ex vivo drug response and clinical outcome.** Overall survival in pediatric BCP-ALL patients from the study cohort, groups colored according to ex vivo drug response (sensitive, intermediate or resistant), p-values calculated using log-rank test. Forest plot showing hazard ratio and 95% confidence interval for relapse in groups with different ex vivo drug response, adjusted for sex and risk group. Results displayed drug wise A-J.
