## Supplementary Figure 2 for "Ex Vivo Drug Responses and Molecular Profiles of 597 Pediatric Acute Lymphoblastic Leukemia Patients"

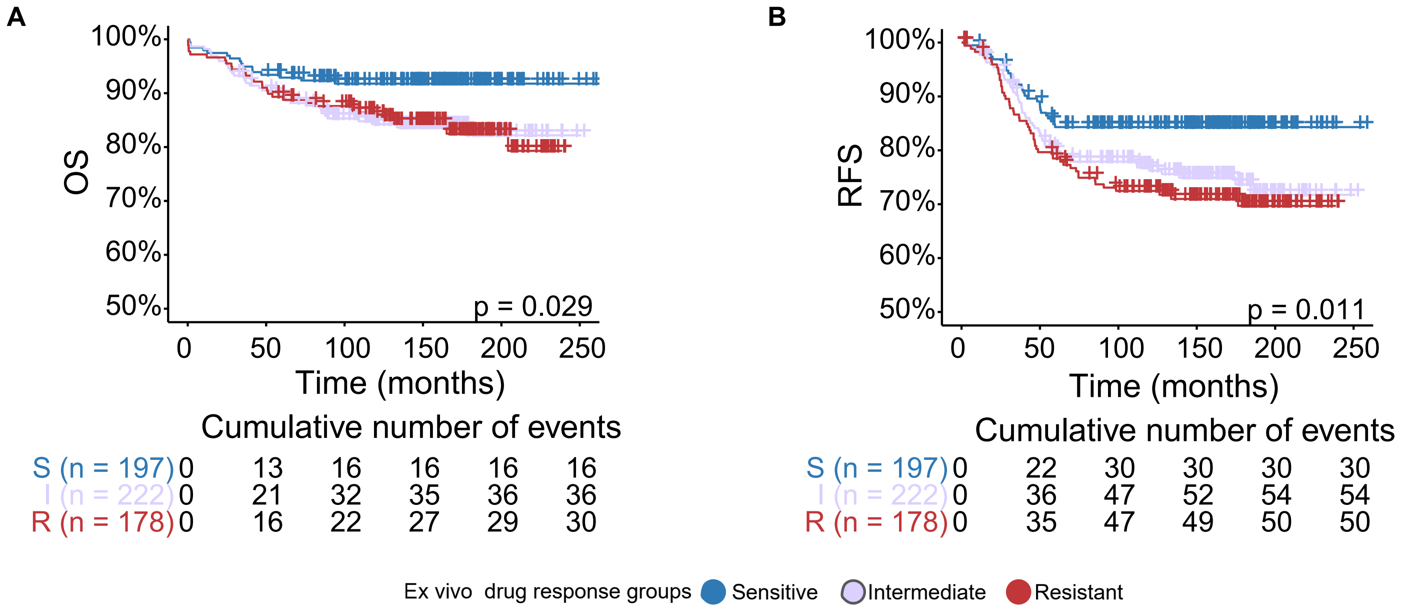


**Supplemental Figure 2. Multidrug response groups have varying clinical outcome and ex vivo drug responses overlap across drugs.** A) Overall survival (OS) and B) relapse free survival (RFS), groups colored according to the different multi-drug response clusters.
