## Supplementary Figure 3 for "Ex Vivo Drug Responses and Molecular Profiles of 597 Pediatric Acute Lymphoblastic Leukemia Patients"

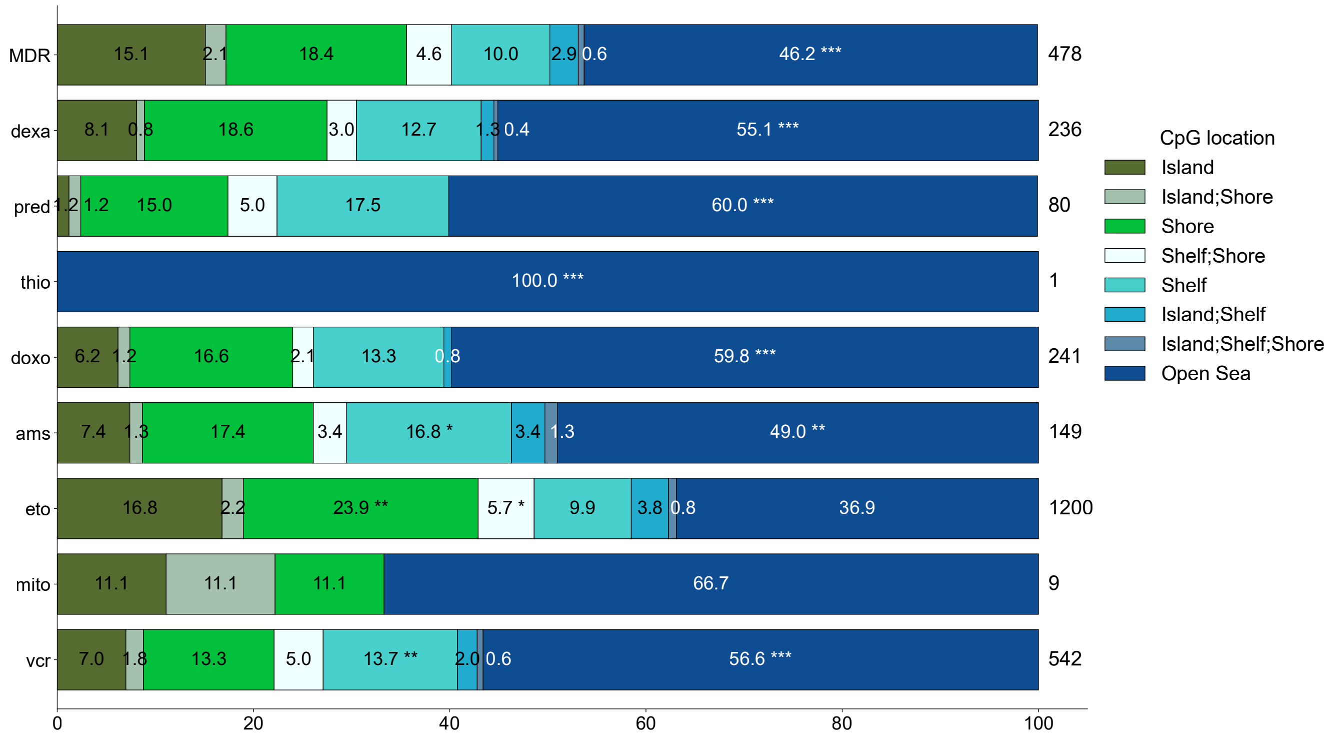


**Supplemental Figure 3. Enrichment of differentially methylated CpG sites to different regions of the genome.** Enrichment of differentially methylated CpG sites to different regions of the genome per drug or in the multidrug resistant cluster (dexa = dexamethasone, pred = prednisolone, cyta = cytarabine, thio = thioguanine, doxo = doxorubicin, ams = amsacrine, eto = etoposide, mito = mitoxantrone, asp = L-asparaginase, vcr = vincristine).
