## Supplementary Methods for "Ex Vivo Drug Responses and Molecular Profiles of 597 Pediatric Acute Lymphoblastic Leukemia Patients"

*Corresponding author:

Dr. Jessica Nordlund

### Supplemental Methods

#### Ex-vivo drug screening

*Ex-vivo* drug responses were assessed by the fluorometric microculture cytotoxicity assay (FMCA).

Diagnostic bone marrow samples were collected in heparinized glass tubes and sent to Uppsala, Sweden for analysis. All samples included in the study contained >70% leukemic blasts and had >70% viability prior to drug screening. FMCA is a method based on measurement of fluorescence generated from the hydrolysis of fluorescein diacetate (FDA) to fluorescein by cells with intact plasma membranes. Each sample was tested for resistance to each drug in triplicate, with six wells without drugs as controls and six wells containing culture medium only as blanks. The mean fluorescence intensity in the drug containing wells (F_test_) was compared to the mean fluorescence intensity in control wells (F_control_) and blank wells (F_blank_) to produce a survival index (SI%):

$$SI\%=100 x \frac{Ftest-Fblank}{Fcontrol-Fblank}$$

#### DNA-methylation arrays and RNA-sequencing

DNA methylation data (DNAm, 450k array) from 382 patients were retrieved from the Gene Expression Omnibus (GEO) entry GSE49031 and <https://figshare.scilifelab.se/> (10.17044/scilifelab.22303531) (1,2). For an additional 55 patients, 15μl of DNA was used and evaporated to a volume of <4μl to determine the genome wide DNA methylation levels using the Infinium MethylationEPIC v2.0 BeadChip assay (EPIC array, Illumina). The raw IDAT files were processed with Illumina GenomeStudio 2011.1 to produce a β-value matrix. All β-values with detection p-value ≥0.01 were replaced with NaNs. Peak-based correction was employed to normalize the β-value distribution of Infinium type I and II probes. The 450k and EPIC array data were then merged into one dataset (n = 437) with 372,264 overlapping CpG sites for downstream analysis. No batch effects were observed after the merge.

Gene expression data (GEX, gene count matrix) for 119 patients were retrieved from the GEO entry GSE227832 (1). The raw read counts were aligned to the reference genome version GRCh38.103 resulting 60,666 genes (ENSMBL IDs) as previously described (1). The gene expression data were corrected for batch effects using ComBat-Seq (3), to remove technical variation due to the different library preparation methods (TruSeq, ScriptSeq, RNAaccess). Finally, the data were normalized using Gene Length corrected trimmed mean of M-values (GeTMM), which adjusts for both gene length and library size, and log2 transformed (4).

#### Bioinformatics and statistical analysis

Differential methylation and expression analyses were performed for each drug between the resistant and sensitive patients. For each drug, CpG sites with > 10% missing values across all the samples were filtered out. Additionally, the β-values were transformed into M-values as presented in (1). M-values produce more robust results in differential methylation analyses with packages such as limma (5).

$M=log2(\frac{beta}{1-beta})$ (1)

Additionally, we adjusted for ALL molecular subtypes to avoid subtype-related biases. The average (mean) β-value difference between the resistant and sensitive patients, which were calculated separately and the adjusted p-values from the differential methylation analyses were collectively used to select the significantly differentially methylated CpG sites for each drug.

For GEX, the differential expression analysis required gene counts as input rather than log2 expression values. Hence, batch corrected counts were introduced to the analysis. Lowly expressed genes (median expression ≤ 5 counts) were filtered out to facilitate the computational time of the analysis resulting in 15,019 genes for downstream analysis. Due to the small sample size for some of the ALL subtypes, we grouped the patients with ≤5 patients into the rest group, before adjusting for molecular subtypes. The DeSeq2 library (6) first normalized the data internally and then produced the average log2 fold change (log2FC) values for the resistant vs the sensitive ex-vivo groups, which combined with the adjusted p-values determined the significantly differentially expresses genes per drug.

DMCs annotation to genes was conducted with the help of GREAT 4.0.4 (7). This tool assigned the DMCs to adjacent genes, enabling the annotation of DMCs located in non-coding regions. For each drug, bed files for the test and background DMCs containing the start and stop coordinates of each DMC, as well their chromosomal position was given as input and was analyzed against GRCh38 reference genome. The annotations were then exported and used for downstream analysis. The drug-specific DMCs and background CpG sites (CpG sites identified as related to drug resistance before the differential methylation analysis) were annotated to locations using Relation_to_UCSC_CpG_Island column from the Illumina EPIC-8v2-0_A1 annotation file. These locations include CpG islands, shores, shelves, and open sea regions. One proportions Z tests followed by Benjamini-Hochberg multiple testing correction (FDR) were performed to detect significantly enriched DMCs in these genomic locations in contrast to the background methylation sites.

In the gene set analysis, the genes used for downstream analysis (n = 15,019) were introduced as a background argument to the pathway analysis to prevent using all the genes from the specified gene sets as background, while computing the statistical significance for each enriched pathway. For the DMC-specific pathway analysis, GREAT gene annotations were used as input. No background was used to perform the analysis, as the majority of the background genes covered the whole genome background. An FDR adjusted p-value of <0.05 determined if the DEGs/DMCs were significantly enriched to a pathway.
